# Aged-black garlic (Abg10+®) supplementation effect in blood lipoprotein profile in Grade I Hypertensive subjects. A randomized, triple-blind controlled trial

**DOI:** 10.64898/2026.04.20.26351262

**Authors:** José CE Serrano, Eva Castro-Boqué, Alicia García-Carrasco, María Inés Morán-Valero, Marina Díez Municio, Marcelino Bermúdez-López, José Manuel Valdivielso, Alberto E Espinel, Manuel Portero-Otín

**Author notes:** Correspondence: Jose CE Serrano, Avenida Alcalde Rovira Roure, 80. Edificio de Biomedicina I, 25198, Lleida, Spain.

## Abstract

Aged black garlic (ABG) is rich in organosulfur compounds such as S⍰allyl⍰cysteine (SAC) and may influence lipid metabolism, although evidence from controlled trials remains limited. The aim of this study was to evaluate the impact of a low-dose SAC-optimized ABG on blood lipid profiles in Grade I hypertensive individuals. A randomized, triple-blind, placebo-controlled trial was conducted with 75 Grade I hypertensive participants receiving either 250 mg of ABG or a placebo daily for 12 weeks. Plasma lipoproteins and subclass composition were quantified by NMR spectroscopy, and metabolic clusters were explored using k⍰means and PLS⍰DA. ABG supplementation led to a significant reduction in the total number of particles and HDL particles. Detailed analysis of XXL-VLDL particles showed a significant decrease in the percentage of both free and esterified cholesterol, alongside an increase in triglyceride percentage. Conversely, large HDL particles exhibited a beneficial remodeling characterized by an increase in phospholipid content and a decrease in triglyceride percentage. Furthermore, cluster analysis demonstrated that participants with a more adverse baseline metabolic profile experienced a significant reduction in total triglycerides and VLDL-lipid content after ABG intake. These results suggest that low-dose ABG supplementation induces specific qualitative improvements in lipoprotein subclasses, particularly enhancing HDL functionality markers and reducing the cholesterol load in large VLDL particles, which may provide cardiovascular benefits in hypertensive individuals with metabolic impairment

## 1. Introduction

Garlic (Allium sativum) has been used for centuries in traditional medicine for its health benefits. Ancient civilizations like the Egyptians, Greeks, Romans and in traditional Chinese and Ayurvedic medicine considered it as a formidable prophylactic and therapeutic medicinal agent (Rahman, 2001). Because of the widespread effects of garlic in maintaining good health, it has attracted particular attention of modern medicine. The effect of garlic in dyslipidemia has been suggested several years ago (Warshafsky, et al., 1993). A recent meta-analysis has found at least 142 articles related to garlic and dyslipidemia (Du et al., 2024) in which it is suggested a significant reduction in Total-cholesterol (TC), Low-density-lipoprotein (LDL) cholesterol and triacylglycerides (TG) blood levels. This is due to its composition of bioactive compounds mainly phenolic and organosulfur compounds (Santhosha et al., 2013), that has been proposed to reduce hepatic lipogenesis and cholesterolegenic enzymes (Yeh & Liu, 2001); enhanced excretion of acidic and neutral steroid into bile salts (Chi, 1982) and suppressed LDL oxidation (Bordia et al., 1998).

However, one of the main problems with these findings is that its translation to clinical practice could be damped since the required doses of garlic to observe the desired effects are relatively high and undesirable symptoms may be observed. Aged Black Garlic (ABG) has gained popularity over regular garlic as a therapeutic treatment due to its enhanced bioavailability, milder taste, and increased antioxidant properties. The fermentation process that transforms fresh garlic into ABG reduces its pungent odor and harsh flavor, making it more appealing for consumption (Kilic-Buyukkurt et al., 2023). Additionally, this process increases the concentration of bioactive compounds such as S-allyl-cysteine (SAC), which has been linked to improved cardiovascular health, anti-inflammatory effects, and stronger antioxidant activity (Shang et al., 2019). Unlike raw garlic, ABG is also easier to digest, reducing the risk of gastrointestinal discomfort. These factors have contributed to its growing use in alternative medicine and functional foods (Ahmed & Wang, 2021).

Several scientific studies have compared the effects of ABG and raw garlic on blood lipid and cholesterol profiles. Research suggests that both types of garlic can reduce TC, LDL cholesterol, and TG, while increasing high-density-lipoprotein (HDL) cholesterol (von Känel-Cordoba et al., 2024). However, ABG appears to have a more potent effect due to its higher concentration of bioactive compounds such as SAC and polyphenols (García-Villalón et al., 2016).

Nevertheless, while traditional lipid profiles focus on TC, LDL-cholesterol, HDL-cholesterol, and TG; studying lipoprotein subclasses provides a more detailed and accurate assessment of cardiovascular risk. LDL is not a single entity; small, dense LDL (sdLDL) particles are more atherogenic than larger, more buoyant LDL particles. Small LDL particles penetrate the arterial wall more easily and are more prone to oxidation, increasing the risk of plaque formation (Borén et al., 2020). Studies show that individuals with high sdLDL levels have a greater risk of CVD even if their overall LDL cholesterol levels appear normal (Hoogeveen et al., 2014). Similarly, not all HDL particles are equally protective. Large HDL particles are more effective in reverse cholesterol transport, whereas small HDL particles may be dysfunctional (Ouimet et al., 2019). In the same context, low HDL particle concentration and dysfunctional HDL composition have been associated with increased cardiovascular risk, independent of total HDL-cholesterol levels (Costacou et al., 2024). Last, smaller VLDL particles are more readily converted into LDL, particularly sdLDL, which is highly atherogenic. Elevated levels of sdLDL are associated with endothelial dysfunction, increased oxidative stress, and enhanced arterial plaque formation, all of which contribute to the progression of atherosclerosis and cardiovascular events (Vekic et al., 2022). Therefore, analyzing blood lipoprotein particle size provides valuable insight into cardiovascular risk beyond traditional lipid measurements.

Hypertension represents a key clinical context in which examining lipid and lipoprotein profiles is particularly relevant. Hypertensive individuals frequently exhibit subtle but clinically meaningful abnormalities in lipid metabolism, even in the absence of overt dyslipidemia (Wang et al., 2024). Studies show that hypertension commonly coexists with increased triglyceride17rich VLDL particles, higher concentrations of small, dense LDL, and reduced levels of large HDL particles—patterns that exacerbate endothelial dysfunction, inflammation, and arterial stiffness (Packard et al., 2020; Paynter et al., 2011; Ramírez-Melo et al., 2025). The coexistence of hypertension and subclinical dyslipoproteinemia amplifies atherosclerotic processes through synergistic metabolic and vascular mechanisms. Therefore, assessing detailed lipid and lipoprotein traits in hypertensive subjects provides important insight into early cardiometabolic derangements and offers a relevant target population for evaluating functional foods with potential lipid17modulating effects. Even modest improvements in lipoprotein dynamics may yield clinically meaningful benefits in this group.

In this context, the present study aimed to evaluate whether daily supplementation with a low17dose, SAC17optimized ABG extract for 12 weeks can modulate the blood lipid profile and the distribution of lipoprotein subclasses in individuals with Grade I hypertension receiving stable antihypertensive therapy. Given the relevance of lipoprotein particle number, size, and composition as early determinants of cardiometabolic risk, these parameters were selected as the main outcomes of the intervention. In addition, LP17IR and other metabolically related biomarkers were included as complementary variables to provide integrated metabolic context and to explore whether ABG induces coordinated changes across lipoprotein17related pathways. Finally, secondary analyses aimed to determine whether specific clinical or metabolic phenotypes exhibit greater responsiveness to ABG supplementation, acknowledging the heterogeneity typically present in hypertensive populations.

## 2. Methods

### 2.1. Experimental Design

The study was designed as a triple-blind, placebo-controlled, randomized parallel trial to assess whether an optimized aged black garlic (ABG) extract could improve the blood lipid profile in subjects with Grade I hypertension. Volunteers were randomly divided into two groups—Group ABG and Group Placebo—and treated for 12 weeks with the active ingredient or placebo, respectively. The treatment duration was determined based on previous studies reported in various meta-analyses on garlic blood lipoproteins (Du et al., 2024; Varade et al., 2024; Zhao et al., 2024).

Volunteers were randomly assigned to each group through sequential number allocation and were stratified by sex after agreeing to participate in the study. The personnel who enrolled and those who assigned participants to the interview had no access to the allocation sequence. The study was conducted in accordance with the ethical principles outlined in the latest versions of the Declaration of Helsinki and Tokyo for human research, as well as Regulation (EU) 2016/679 of the European Parliament and Council of April 27, 2016, on Data Protection (GDPR); Law 14/2007 on biomedical research; and Organic Law 3/2018 on the Protection of Personal Data and Guarantee of Digital Rights. Participants provided informed consent before joining the study. The study protocol was approved by the Hospital Universitari Arnau de Vilanova (CEIC-2435), Lleida, Spain, and the trial is registered at ClinicalTrials.gov under the identifier NCT04915053. This trial was designed and reported in accordance with the CONSORT 2010 Statement for randomized controlled trials. No changes to the methods were introduced after the trial commenced.

### 2.2. Intervention

The intervention consisted of the daily intake of one tablet upon waking and before breakfast. The ABG group received a tablet of optimized aged black garlic extract (Abg10+®/Garlace®), standardized in Agedgarlides® (a totum of the main organosulfur compounds found in aged garlic, responsible for the mechanism of action and the scientifically proven bioactivity) and manufactured by Pharmactive Biotech Products, S.L.U. (Madrid, Spain) through ActiveNatureTM Technology. The extract was derived from garlic grown in the Las Pedroñeras region (Castilla-La Mancha, Spain), and the tablets were produced by Instant Procès (La Roca del Vallès, Spain). Each ABG tablet contained 250 mg of ABG extract, equivalent to 0.25 mg of SAC per tablet, along with 300 mg of excipients. Detailed composition is described in **Table 1**. The placebo group received a tablet in which the ABG extract was replaced with maltodextrin. Both tablets were similar in appearance and odor and were coded by the manufacturer. To ensure blinding, none of the researchers involved in the study knew to which group each tablet had been assigned. Participants were instructed to maintain their dietary habits, physical activity, and medical treatments. Adverse events were asked systematically in each interview. No interim analysis was performed. At the end of the experimental phase, any changes in lifestyle and pharmacological treatment were assessed, with no volunteers reporting modifications. Volunteer recruitment and the start of the trial began in September 2021 and concluded in April 2022.

**Table 1.**
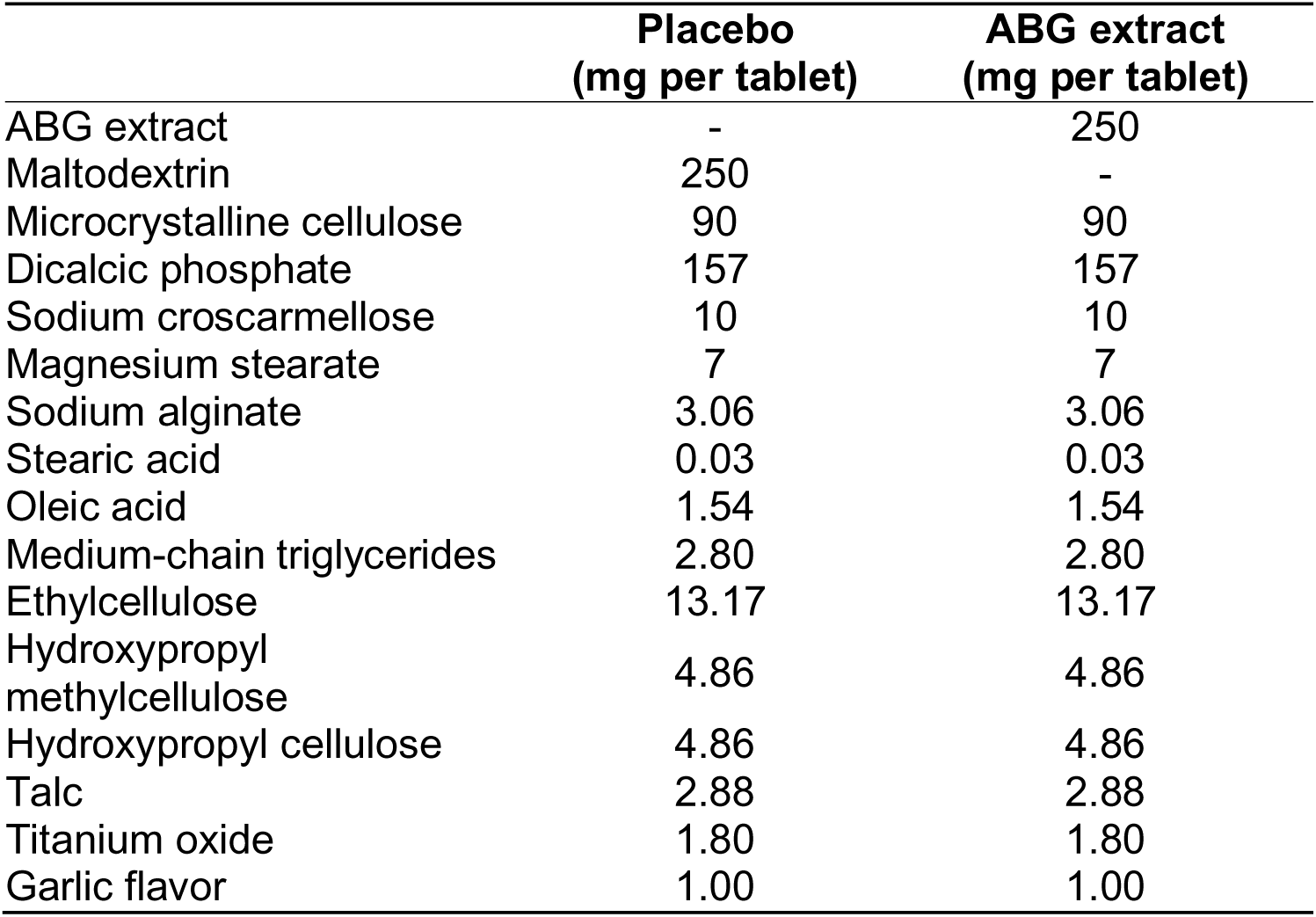
Detailed composition of tablets employed in the study.

### 2.3. Study Population

Volunteers were recruited in collaboration with the Atherothrombotic Disease Detection and Treatment Unit (UDETMA) at the Hospital Universitari Arnau de Vilanova (Lleida, Spain) from a database of patients diagnosed with Grade I hypertension. The inclusion criteria encompassed men and women over 18 years old with hypertension under treatment with one or more antihypertensive drugs, excluding Angiotensin-Converting-Enzyme inhibitors. Exclusion criteria included morbid obesity (body mass index > 35 kg/m²), serum LDL-cholesterol levels below 115 mg/dL (threshold for low or moderate SCORE risk of cardiovascular events), fasting serum glucose levels above 126 mg/dL (cut-off for diabetes diagnosis), anemia (hemoglobin < 13 g/dL in men and < 12 g/dL in women), chronic gastrointestinal disease, pregnancy or intention to become pregnant, lactation, participation or planned participation in a clinical trial or nutritional intervention study within the 30 days prior to enrolment, and allergy to garlic.

### 2.4. Sample Size Determination

The sample size was estimated based on available evidence regarding the variability of traditional lipid parameters and the expected lipid17lowering effects of garlic-derived preparations. Within17person variability for total cholesterol has been reported to be low (SD ≈ 0.12 mmol/L) (Pereira et al., 2004), and meta17analytic data suggest that garlic supplementation may reduce total cholesterol by approximately 0.37 mmol/L (Zhao et al., 2024). Under these assumptions, a sample size of approximately 36 participants per group provides very high statistical power to detect changes of this magnitude using a two17sided α = 0.05. In addition to traditional lipid measures, the present study incorporated NMR17derived lipoprotein subclasses and LP17IR as complementary metabolic variables. These advanced parameters typically exhibit greater biological variability and smaller expected effect sizes than routine lipids. Therefore, although the study was adequately powered for detecting moderate changes in conventional lipid markers, it was not specifically powered to detect small or subtle modifications in lipoprotein particle composition. Findings related to these exploratory outcomes should thus be interpreted as hypothesis17generating.

### 2.5. Outcomes

#### 2.5.1. Blood lipid profile

Lipid profiles were investigated by the Nightingale Health’s NMR spectroscopy method (Deelen et al., 2019; Holmes et al., 2018; Würtz et al., 2017). Following the blood draw, blood samples were stored at −80 °C prior to shipment to the Metabolomics Core Facility at the University of Bristol. 250 biomarkers were determined which included routine lipids, apolipoproteins, amino acids, fatty acids, lipoprotein particles, inflammation and glycolysis-related metabolites.

#### 2.5.2 Lipoprotein Insulin Resistance Index (LP-IR)

LP-IR was calculated from NMR-derived lipoprotein parameters (Shalaurova et al., 2014). The index integrates six lipoprotein subclass features associated with insulin17resistant dyslipidemia: large VLDL particle concentration, small LDL particle concentration, large HDL particle concentration, and the mean particle sizes of VLDL, LDL, and HDL. For each participant, LP⍰IR was computed on the 0–100 scale, where higher values indicate a more insulin17resistant lipoprotein pattern.

#### 2.5.3 Anthropometric measurements

Anthropometric measurements were obtained following standardized procedures. Body weight was measured to the nearest 0.1 kg using a calibrated scale, with participants wearing light clothing and no shoes. Height was measured to the nearest 0.1 cm using a wall⍰mounted stadiometer, ensuring an upright posture with heels, buttocks, and upper back aligned. Body mass index (BMI) was calculated as weight divided by height squared (kg/m²). Waist and hip circumferences were assessed using a non⍰elastic tape placed horizontally at standardized anatomical landmarks. Waist circumference was measured midway between the lowest rib and the iliac crest after gentle exhalation, and hip circumference at the level of the greatest gluteal protrusion. Two measurements were taken for each parameter, and a third measurement was performed when the first two differed by more than 0.5 cm. The waist⍰to⍰hip ratio was calculated as waist circumference divided by hip circumference.

#### 2.5.4. Metabolic syndrome classification

Metabolic syndrome classification was done according to the Adult Treatment Panel III (ATP III) of the National Cholesterol Education Program (NCEP) guidelines (undefined, 2002). Metabolic Syndrome was classify on the presence of at least three out of five criteria that include: abdominal obesity (waist circumference >102 cm in men and >88 cm in women), elevated triglycerides (≥150 mg/dL or drug treatment for high triglycerides), low HDL cholesterol (<40 mg/dL in men and <50 mg/dL in women or drug treatment for low HDL), high blood pressure (≥130/85 mmHg or use of antihypertensive medication), and elevated fasting glucose (≥100 mg/dL or current treatment for hyperglycemia).

#### 2.5.5. Endothelial Function

Endothelial function was evaluated using the EndoPAT™ parameters, including the reactive hyperemia index (RHI) and the augmentation index (AI_75_), both assessed at baseline and after 12 weeks of intervention. Analyses were conducted in a room maintained at a thermoneutral temperature of 24 °C. Volunteers were instructed to remove any restrictive clothing, watches, or rings that could hinder blood flow to the arms and fingers. The blood pressure cuff was then snugly placed on the upper arm. Participants lay comfortably in the study room, relaxing for at least 15 minutes before the test began. The measurement protocol comprised 1 minute of initial reading to verify proper sensor placement and signal quality, followed by 5 minutes under basal conditions, 5 minutes of occlusion, and 5 minutes of recording post-occlusion. Occlusion was achieved by inflating the cuff to a supra-systolic level, at least 60 mmHg above the systolic blood pressure and not below 200 mmHg. Blood-flow cessation was confirmed by the total absence of signal from the occluded hand. If any movement was detected, cuff pressure was increased by an additional 50 mmHg, up to a maximum of 300 mmHg. The EndoPAT™ 2000 software automatically calculated RHI and AI_75_ values. Occlusion parameters were reviewed for all measurements, with manual adjustments made as needed.

#### 2.5.6. Blood Pressure

Blood pressure measurements were obtained using the same oscillometric automatic sphygmomanometers (Medisana BU 512, Medisana GmbH, Neuss, Germany), with a maximum static pressure error tolerance of ±3 mmHg. Volunteers remained seated comfortably in a quiet environment for 10 minutes before the measurements. Blood pressure values were determined by averaging three consecutive readings of systolic and diastolic blood pressure, taken at 3-minute intervals while participants were seated with their feet flat on the floor and arms supported at heart level.

### 2.6. Statistical Analysis

Primary and secondary parameters were measured at baseline and after 12 weeks of intervention. All the data was included in the statistical analysis, and no missing data was observed. Data are presented as mean ± standard deviation of the mean. Two-way ANOVA and multiple comparisons, used to analyze the treatment effect, were performed using Sidak’s test. A statistical analysis of initial differences between Groups ABG and placebo was performed by a two-tailed unpaired Student’s t-test. A Chi-square test was performed for categorical variables.

A multivariable linear regression model was used to evaluate the association between the change in blood pressure and lipoprotein subclasess. The model included an interaction term between change in lipoproteins and study group (Placebo vs. ABG) to assess whether the association differed between groups. The analysis was adjusted for age, sex, body mass index (BMI), and previous lipid-lowering medication. A two-sided p-value <0.05 was considered statistically significant.

To identify distinct patterns within lipid profile data, k-means clustering using the MetaboAnalyst 6.0 platform (https://www.metaboanalyst.ca/) was employed. It was specified a k = 3, resulting in the formation of three distinct clusters. Further to identify the distinguishing features of each cluster, Partial Least Squares Discriminant Analysis (PLS-DA) using the MetaboAnalyst platform was applied. To assess the statistical differences of each variable across clusters, volcano plots within the MetaboAnalyst platform were performed. Finally, in all cases, the cut-off to determine significant differences between groups was set at a p-value below 0.05. Statistical analyses and graphs were achieved with GraphPad Prism v9 and MetaboAnalyst platform.

## 3. Results

### 3.1. Study design

A flow chart of the study design and volunteer included in each phase is depicted in **Figure 1**. A total of 303 patients who met the inclusion and exclusion criteria were contacted; of these, 89 expressed interests in participating in the study, 165 were not interested, and 49 did not answer the phone call. The 89 patients interested in participating in the study were randomly assigned to two groups: 44 volunteers in the Placebo Group and 45 volunteers in the ABG Group. All of them were scheduled for an initial appointment to provide study instructions. Of the 89 volunteers scheduled, 8 did not attend (4 from the Placebo group and 4 from the ABG group) for the first appointment, leaving 81 volunteers who started the experimental phase. At the end of the experimental phase, 4 volunteers from the placebo group and 1 volunteer from the ABG group were excluded due to loss to follow up. Last, 1 volunteer from the ABG group discontinued the study due to treatment intolerance. Finally, the data and sample analysis of a total of 36 and 39 volunteers from the Placebo and ABG groups, respectively, were analyzed and included in the study results. After that, no data or samples were excluded from the final analysis. The baseline characteristics of the volunteers are described in **Table 2**. No significant differences were observed in the initial lipid profile parameters between the Placebo and ABG groups.

**Figure 1.**
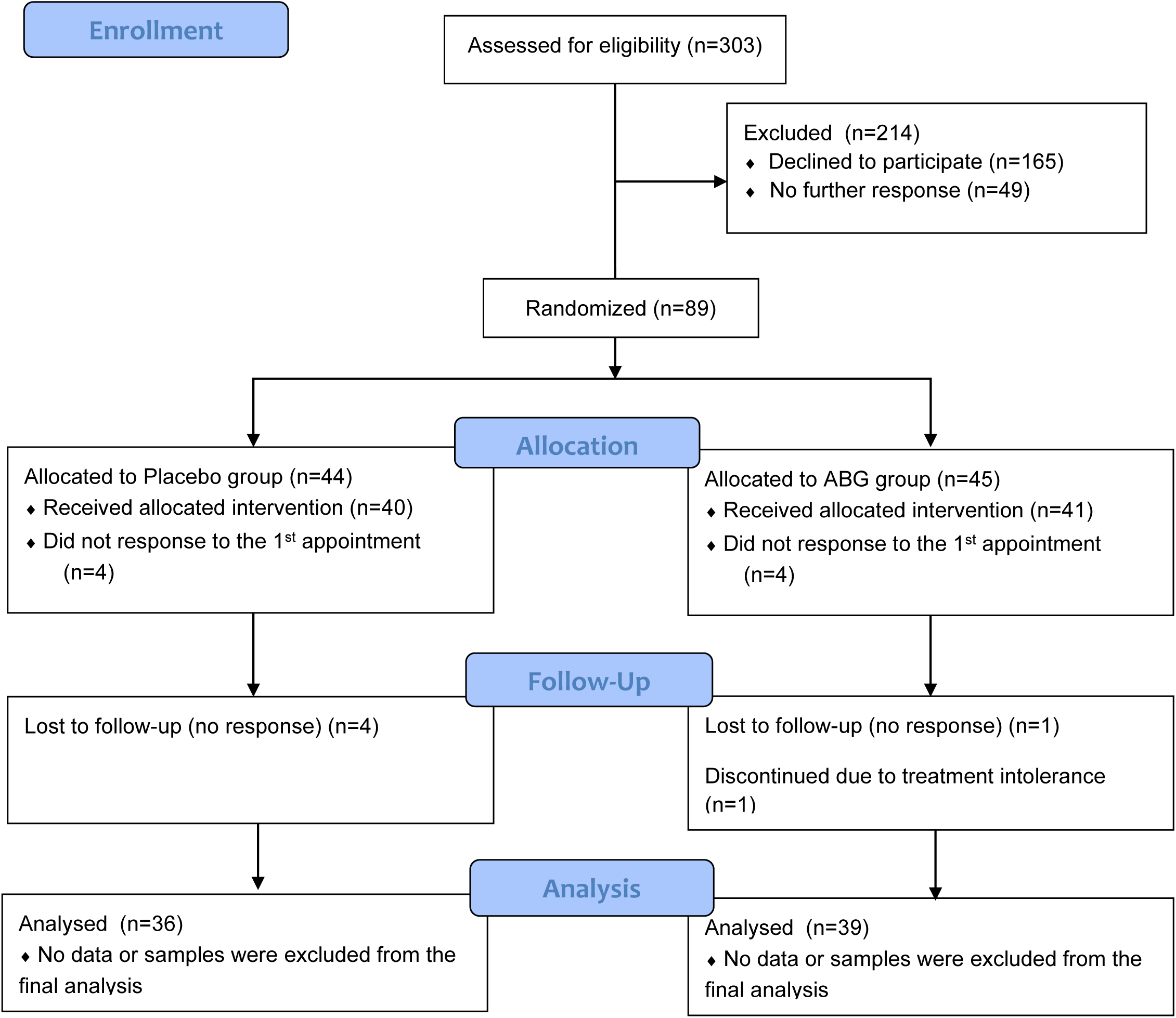
Flow chart of study design and volunteers’ distribution and dropouts in each phase.

**Table 2.** Volunteer’s basal characteristics. Data is presented as mean ± standard deviation. Differences between groups were determined by two-tailed t-student for parametric variables or chi-squared for non-parametric variables. P-value below 0.05 were considered as statistical differences between groups.

| Variable | Group Placebo | Group ABG | p-value |
| --- | --- | --- | --- |
| Number of volunteers | 36 | 39 |  |
| Sex (male:female) | 20:16 | 20:19 | 0.8178 |
| Body mass index (kg/m<sup>2</sup>) | 31.9 ± 5.4 | 30.9 ± 4.9 | 0.4011 |
| Waist/hip ratio |  |  |  |
| Men | 1.02 ± 0.06 | 1.00 ± 0.07 | 0.4792 |
| Women | 0.90 ± 0.06 | 0.93 ± 0.07 | 0.1627 |
| Lipid lowering treatment (yes/no) | 20/16 | 16/23 | 0.2513 |
| Blood glucose (mg/dL) | 108 ± 16 | 106 ± 12 | 0.5547 |
| Total blood lipids (mmol/L) | 9.95 ± 1.74 | 9.23 ± 1.88 | 0.0883 |
| Blood triacylglycerides (mmol/L) | 1.46 ± 0.66 | 1.30 ± 0.66 | 0.2796 |
| Total blood cholesterol (mmol/L) | 5.33 ± 1.00 | 4.94 ± 0.97 | 0.0927 |
| HDL-Cholesterol (mmol/L) | 1.44 ± 0.40 | 1.41 ± 0.36 | 0.7011 |
| LDL-Cholesterol (mmol/L) | 2.25 ± 0.52 | 2.07 ± 0.48 | 0.1175 |
| VLDL-Cholesterol (mmol/L) | 0.71 ± 0.26 | 0.62 ± 0.26 | 0.1525 |
| LP-IR score | 39.6 ± 4.4 | 39.9 ± 4.4 | 0.8014 |

### 3.2. Treatment Adherence and Reported Adverse Events

Treatment compliance was calculated by dividing the number of pills taken by the total number of treatment days. Some volunteers extended their treatment period due to scheduling adjustments for appointments. The average number of treatment days was 87 ± 5 in ABG group and 88 ± 5 in Placebo group. Both groups showed a mean compliance rate above 96%, with no significant difference between the groups (p = 0.4396). All data was retained for statistical analysis. Adverse events were reported by five volunteers and did not interfere with treatment adherence. Two participants in the ABG group and two in the placebo group experienced mild gastric discomfort, while one additional participant in the placebo group reported muscle cramps. No severe adverse events were recorded.

To evaluate the blinding process, participants were asked at their final appointment whether they believed they were in the active or placebo group. In response, 23% of volunteers in the ABG group and 45% in the Placebo group believed they were in the active group, confirming that blinding was effective. The researchers also remained unaware of group assignments until the completion of the statistical analysis.

### 3.3. Lipid profile

**Table 3** shows the effect of ABG treatment on the lipid profile. After 12 weeks of treatment, no significant change is observed in any of the usually analyzed biomarkers in clinical practice, nor in the LP-IR score and TG/HDL-cholesterol ratios. Similarly, it was analyzed whether the response to treatment differs based on the sex of the volunteer and whether they receive medication to reduce cholesterol (**Supplementary information, Tables S1 and S2**). It was observed that men significantly modified their total cholesterol levels, without observing differences in terms of the treatment received. Regarding the effect of lipid-lowering treatment, 30 volunteers reported the use of statins (13 Atorvastatin, 16 Simvastatin, and 1 Pravastatin) and one volunteer reported the use of Ezetimibe. It was observed that lipid-lowering treatment does not have any influence on the response to treatment with ABG.

**Table 3.**
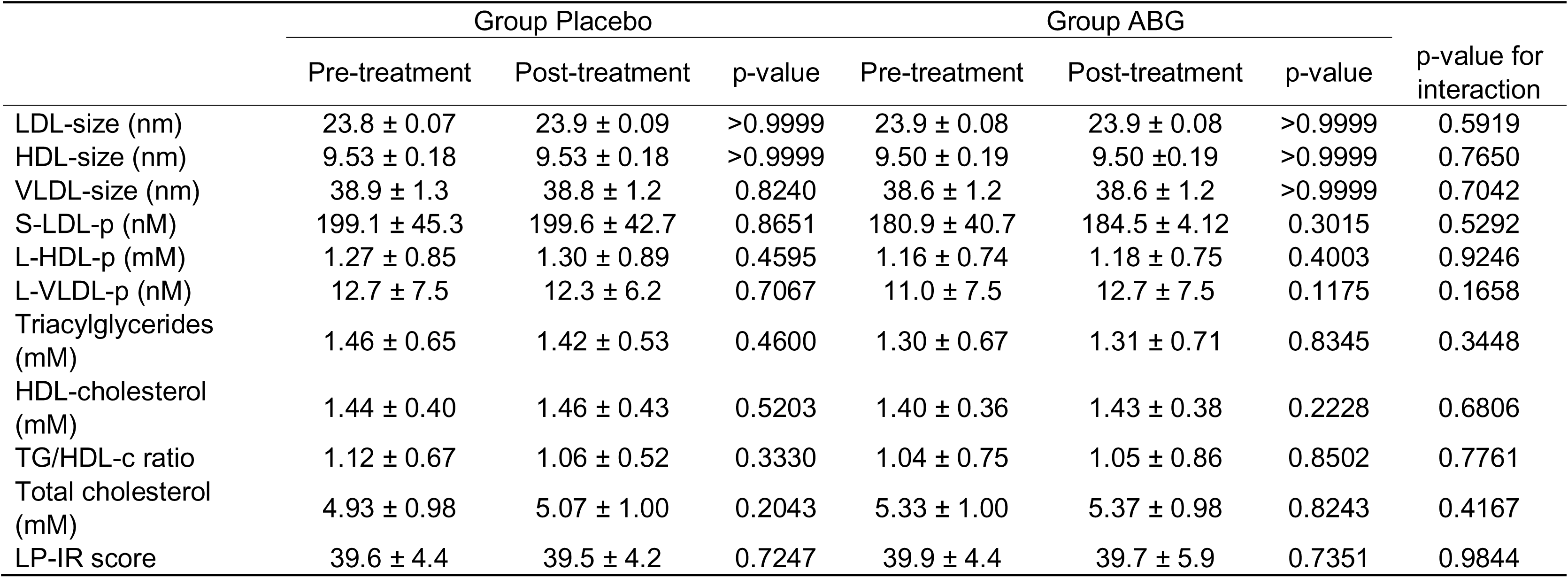
Changes in lipoprotein subclasses related to metabolic syndrome. Data is presented as mean ± standard deviation. Differences between groups was determined by two-way ANOVA. Bonferroni’s multiple comparison test was used to determine differences by time and group treatment. P-value below 0.05 were considered as statistical differences between groups.

A detailed analysis of the lipoprotein profile is shown in **Figure S1** (Supplementary information), where the 95% confidence intervals of the difference between the Placebo and ABG groups after 12 weeks of treatment are depicted. Similarly, in **Table 4** differences and 95% confidence intervals of the differences of the features that presented a statistically significant change are described. A decrease in the total number of particles (Total-P) (p=0.0136), primarily in HDL particles (p=0.0252) in the ABG treated group was observed. Differences were also present in the composition of XXL-VLDL, where the ABG group showed a higher percentage of triglycerides (p<0.0001) and a lower percentage of free and esterified cholesterol (p<0.0001 in both) in XXL-VLDL particles. In contrast, in L-HDL particles the ABG group had a lower percentage of triglycerides (p<0.0036) and a higher percentage of phospholipids (p<0.0277) compared to the placebo.

**Table 4.** Main lipoprotein particles difference between treatments. The data is presented as the main differences between Placebo and ABG+ treatment. Two way ANOVA was used to determine treatment effect. Bonferroni’s multiple comparison test was used to determine differences by time and group treatment. P-value below 0.05 were considered as statistical differences between groups.

| Variable | Mean difference<br>(Placebo – ABG+) | 95% CI of the<br>difference | p-value |
| --- | --- | --- | --- |
| Total particles | $2.7 \times 10^{-4}$ | $5.7 \times 10^{-5}$ to $4.9 \times 10^{-4}$ | 0.0136 |
| HDL-particles | $2.5 \times 10^{-4}$ | $3.1 \times 10^{-5}$ to $4.7 \times 10^{-4}$ | 0.0252 |
| XXL-VLDL-C% | 13.8 | 9.6 to 18.1 | <0.0001 |
| XXL-VLDL-CE% | 13.1 | 8.8 to 17.3 | <0.0001 |
| XXL-VLDL-TG% | -11.28 | -15.5 to -7.1 | <0.0001 |
| L-HDL-PL% | -1.98 | -3.7 to -0.2 | 0.0277 |
| L-HDL-TG% | 2.63 | 0.9 to 4.4 | 0.0036 |

A linear regression model was fitted to evaluate the association between the change in blood pressure and the change in lipoproteins classes, including as covariates previous lipid-lowering medication, sex, age and IMC. It was observed that the change in systolic blood pressure was directly correlated with the change in blood total cholesterol levels (p=0.0166) and LDL-C (p=0.0495). Similarly, it was that the between the change in total cholesterol and group showed a trend toward significance (p=0.0703), suggesting that the relationship between cholesterol and systolic blood pressure may differ between groups (**Figure 2**). Similar analysis were performed with other lipoprotein subclasses, however, none of them showed statistical significance (for example with systolic blood pressure HDL-C (p=0.0969); TG (p=0.977); and diastolic blood pressure Total-cholesterol (p=0.058); LDL-C (p=0.162); HDL-C (p=0.122) and TG (p=0.969).

**Figure 2.**
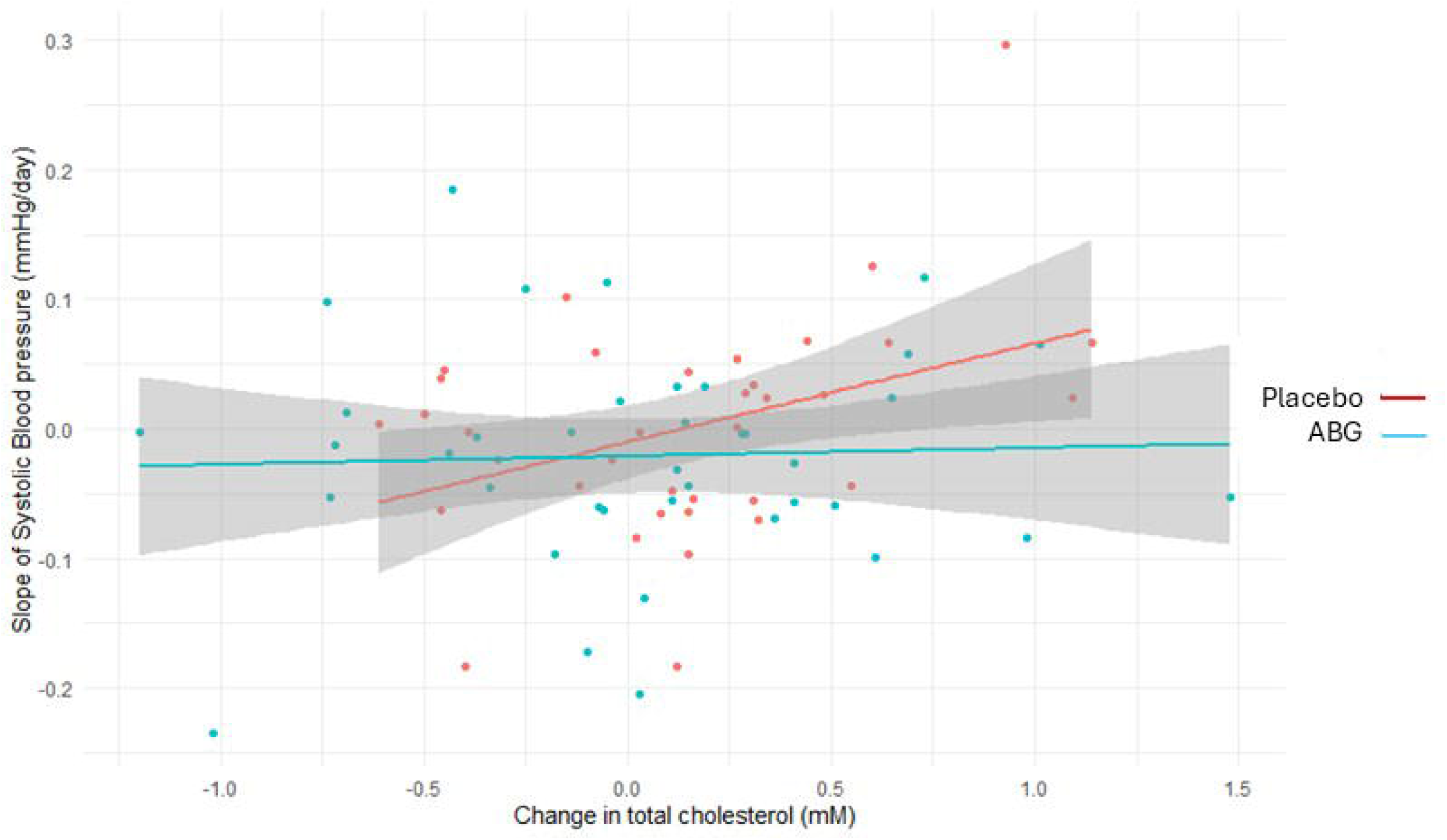
Association between change in total cholesterol and the slope of systolic blood pressure according to study group. Each point represents an individual participant. The x-axis shows the change in total cholesterol (mM), and the y-axis shows the slope of systolic blood pressure (mmHg/day). Linear regression lines are shown separately for the placebo group and the ABG group.

### 3.4. Cluster analysis

Considering that the response to treatment may be influenced by the baseline lipoprotein levels, a cluster analysis was conducted to classify the volunteers based on their lipoprotein profile. Three distinct clusters were identified, each with differential characteristics of metabolic syndrome, as shown in **Table 5**. For example, Cluster 1 shows a lower incidence of metabolic syndrome and is characterized by lower levels of body mass index, blood lipids, LP-IR score and augmentation index (AI_75_). On the opposite end, Cluster 3 is characterized by having more than three components of the metabolic syndrome classification according to NCEP ATP III, which are higher body mass index, high blood glucose, low HDL-cholesterol and high blood triacylglycerides levels, LP-IR score and glycoprotein acetyls a biomarker of inflammation. Main differences in lipoprotein subclasses are described in **Figure 3**. The PLS-DA confirmed that each cluster could be identified by a specific lipid profile, being the main variables in importance that may classify each cluster the XXL-VLDL lipoprotein. In which, Cluster 3 is characterized by XXL-VLDL lipoproteins with high levels of TG, lipids and phospholipids; while, on the other hand, Cluster 1 present lower levels.

**Figure 3.**
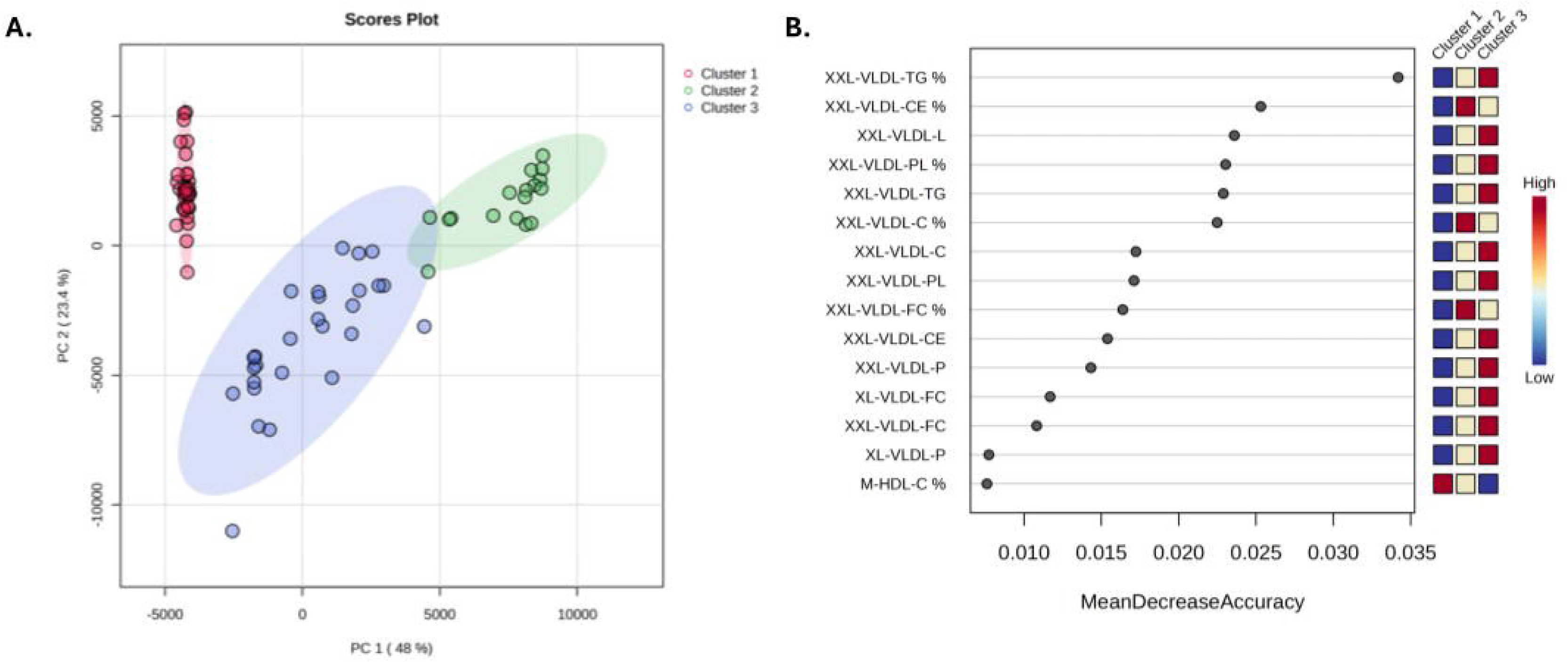
Main lipoprotein subclasses that identify each cluster. A. 2D Score plot of the PLS-DA cluster classification. PLS-DA cross validation for 5 components: Accuracy 0.9875; R2 0.9692; and Q2 0.9590. B. Variable importance in projection (VIP) and weighted sum of absolute regression coefficients. The colored boxes on the right indicate the relative concentrations of the corresponding metabolite in each group under study.

**Table 5.** Clusters demographic characteristics at base line. Data is presented as mean ± standard deviation. Differences between clusters for continuous variables was determined by one-way ANOVA. Bonferroni’s multiple comparison test was used to determine differences by group. For categorical variables a Chi-squared test was performed. P-value below 0.05 were considered as statistical differences between groups. Differences between clusters were denoted with superscript as follows: a denote difference with Cluster 1, and b denote difference with Cluster 2; while *, **, ***, and **** denotes p values of <0.05, <0.01, <0.001 and <0.0001 respectively.

|  | **Cluster 1**<br>**(n=31)** | **Cluster 2**<br>**(n=17)** | **Cluster 3**<br>**(n=27)** | **p-value** |
| --- | --- | --- | --- | --- |
| Metabolic syndrome | 8 (26%) | 6 (35%) | 16 (60%) | 0.0328 |
| Treatment (Placebo:ABG) | 18:13 | 7:10 | 11:16 | 0.3698 |
| Age (years) | 63.6 ± 5.7 | 65.5 ± 4.9 | 62.9 ± 6.2 | 0.3236 |
| Sex (male:female) | 16:15 | 6:11 | 18:9 | 0.1369 |
| Systolic blood pressure (mmHg) | 145.2 ± 18.9 | 144.8 ± 20.3 | 151.2 ± 22.2 | 0.4671 |
| Diastolic blood pressure (mmHg) | 85.7 ± 11.9 | 86.0 ± 9.7 | 85.1 ± 11.1 | 0.9656 |
| Body mass index (kg/m<sup>2</sup>) | 29.5 ± 4.5 | 30.3 ± 4.1 | 34.9 ± 4.8<sup>a****,b**</sup> | <0.0001 |
| RHI | 2.06 ± 0.54 | 1.94 ± 0.36 | 2.08 ± 0.51 | 0.6264 |
| AI<sub>75</sub> | 31.7 ± 13.4 | 31.2 ± 15.8 | 20.7 ± 17.2<sup>a*</sup> | 0.0196 |
| Glycoprotein Acetyls (mM) | 0.85 ± 0.09 | 0.88 ± 0.06 | 0.96 ± 0.12<sup>a***,b*</sup> | 0.0002 |
| Blood glucose (mg/dL) | 107.6 ± 14.5 | 100.3 ± 8.2 | 111.7 ± 16.5<sup>b*</sup> | 0.0426 |
| Blood cholesterol (mM) | 4.77 ± 0.88 | 5.86 ± 1.12<sup>a***</sup> | 5.10 ± 0.84<sup>b*</sup> | 0.0010 |
| HDL-cholesterol (mM) | 1.59 ± 0.43 | 1.47 ± 0.28 | 1.20 ± 0.24<sup>a***,b*</sup> | 0.0002 |
| LDL-cholesterol (mM) | 1.91 ± 0.42 | 2.52 ± 0.56<sup>a***</sup> | 2.21 ± 0.44<sup>a*</sup> | 0.0002 |
| Blood triacylglycerides (mM) | 0.88 ± 0.23 | 1.38 ± 0.26<sup>a**</sup> | 1.95 ± 0.70<sup>a****,b***</sup> | <0.0001 |
| LP-IR score | 36.4 ± 4.3 | 40.6 ± 2.5<sup>a***</sup> | 43.2 ± 2.0a<sup>****,b*</sup> | <0.0001 |

The main effect of the treatment in main lipoprotein classes, according to the cluster to which each volunteer belongs, was a decrease in triglyceride levels (p=0.0154 for volunteers in Cluster 3 treated with ABG and p=0.0316 for the difference between clusters in the ABG treatment). However, it is also observed that volunteers in Cluster 1 treated with ABG show a significant increase in glucose levels (**Table 6**). Considering that the main effect of ABG treatment was a decrease in triglycerides, an analysis of the lipoprotein profile in VLDL lipoproteins was performed. It was observed that in Cluster 3, which is characterized by high levels of XXL-VLDL, ABG treatment reduces the lipid content in these particles (p=0.0155) as well as the TG of VLDL particles (p=0.0131). Additionally, a significant increase in lipid content (p=<0.0001), mainly phospholipids (p=0.0388), in XL-HDL particles were observed.

**Table 6.** Changes in metabolic syndrome parameters derived by treatment and cluster belonging. Data is presented as mean ± standard deviation or mean difference of the change of each parameter (ABG – Placebo) and 95% confidence interval of the difference. Differences between treatments and clusters was determined by two-way ANOVA. Bonferroni’s multiple comparison test was used to determine differences by treatment/cluster. P-values below 0.05 was considered as statistical difference.

|  | Cluster 1 |  |  | Cluster 2 |  |  | Cluster 3 |  |  | p-value for Cluster |
| --- | --- | --- | --- | --- | --- | --- | --- | --- | --- | --- |
|  | Basal | 12 weeks | p-value | Basal | 12 weeks | p-value | Basal | 12 weeks | p-value |  |
| **Group Placebo** |  | n=18 |  |  | n=7 |  |  | n=11 |  |  |
| Systolic blood pressure (mmHg) | 144 ± 16 | 140 ± 13 | 0.3054 | 144 ± 12 | 139 ± 12 | 0.4246 | 149 ± 22 | 140 ± 14 | 0.0523 | 0.6602 |
| Diastolic blood pressure (mmHg) | 85 ± 12 | 85 ± 11 | 0.7799 | 84 ± 3 | 83 ± 10 | 0.8227 | 85 ± 13 | 82 ± 9 | 0.3080 | 0.8117 |
| Glycoprotein acetyls (mM) | 0.87 ± 0.09 | 0.85 ± 0.08 | 0.6981 | 0.86 ± 0.06 | 0.90 ± 0.07 | 0.5081 | 0.98 ± 0.14 | 0.98 ± 0.12 | 0.9793 | 0.7440 |
| Blood glucose (mg/dL) | 109 ± 15 | 103 ± 15 | 0.0058 | 102 ± 10 | 103 ± 8 | 0.6548 | 107 ± 11 | 110 ± 17 | 0.8026 | 0.0626 |
| Blood cholesterol (mM) | 4.62 ± 0.75 | 4.76 ± 0.87 | 0.1872 | 5.49 ± 1.17 | 5.59 ± 1.12 | 0.5588 | 5.09 ± 1.09 | 5.24 ± 1.04 | 0.2581 | 0.9663 |
| HDL-cholesterol (mM) | 1.53 ± 0.33 | 1.57 ± 0.38 | 0.2526 | 1.39 ± 0.39 | 1.38 ± 0.36 | 0.8867 | 1.20 ± 0.32 | 1.23 ± 0.34 | 0.4153 | 0.7508 |
| LDL-cholesterol (mM) | 1.87 ± 0.37 | 1.92 ± 0.37 | 0.3905 | 2.35 ± 0.43 | 2.39 ± 0.46 | 0.6636 | 2.22 ± 0.57 | 2.31 ± 0.51 | 0.1941 | 0.8483 |
| Triacylglycerides (mM) | 0.86 ± 0.25 | 0.88 ± 0.18 | 0.7824 | 1.31 ± 0.24 | 1.42 ± 0.29 | 0.2848 | 2.00 ± 0.73 | 1.95 ± 0.93 | 0.4847 | 0.4413 |
| LP-IR score | 36.7 ± 3.9 | 36.8 ± 3.8 | 0.6610 | 41.3 ± 2.1 | 41.4 ± 1.6 | 0.8145 | 43.5 ± 2.6 | 42.5 ± 3.0 | 0.0680 | 0.1965 |
| **Group ABG** |  | n=13 |  |  | n=10 |  |  | n=16 |  |  |
| Systolic blood pressure (mmHg) | 147 ± 23 | 142 ± 17 | 0.1500 | 147 ± 25 | 139 ± 18 | 0.1085 | 153 ± 23 | 147 ± 20 | 0.0885 | 0.9754 |
| Diastolic blood pressure (mmHg) | 86 ± 12 | 84 ± 7 | 0.3636 | 87 ± 12 | 84 ± 8 | 0.1802 | 85 ± 10 | 83 ± 10 | 0.3970 | 0.8908 |
| Glycoprotein acetyls (mM) | 0.82 ± 0.09 | 0.86 ± 0.11 | 0.4018 | 0.89 ± 0.06 | 0.92 ± 0.10 | 0.4409 | 0.94 ± 0.10 | 0.92 ± 0.08 | 0.4177 | 0.4015 |
| Blood glucose (mg/dL) | 105 ± 15 | 109 ± 19 | 0.3279 | 99 ± 7 | 103 ± 25 | 0.3657 | 115 ± 19 | 110 ± 20 | 0.2569 | 0.2331 |
| Blood cholesterol (mM) | 4.99 ± 1.02 | 5.06 ± 1.12 | 0.6600 | 6.12 ± 1.07 | 6.10 ± 0.97 | 0.9269 | 5.11 ± 0.66 | 5.17 ± 0.60 | 0.6698 | 0.9255 |
| HDL-cholesterol (mM) | 1.67 ± 0.56 | 1.65 ± 0.65 | 0.5208 | 1.54 ± 0.17 | 1.54 ± 0.21 | 0.9808 | 1.21 ± 0.16 | 1.26 ± 0.18 | 0.1248 | 0.3021 |
| LDL-cholesterol (mM) | 1.98 ± 0.48 | 2.01 ± 0.50 | 0.5208 | 2.64 ± 0.60 | 2.63 ± 0.58 | 0.8923 | 2.22 ± 0.34 | 2.24 ± 0.31 | 0.7670 | 0.9481 |
| Triacylglycerides (mM) | 0.91 ± 0.22 | 1.02 ± 0.23 | 0.3108 | 1.44 ± 0.28 | 1.53 ± 0.55 | 0.4697 | 1.92 ± 0.70 | 1.67 ± 0.52 | 0.0154 | 0.0316 |
| LP-IR score | 36.0 ± 4.9 | 36.5 ± 8.3 | 0.5986 | 40.0 ± 2.8 | 39.7 ± 4.7 | 0.7967 | 43.0 ± 1.5 | 42.4 ± 2.1 | 0.4984 | 0.6906 |
| **ABG vs Placebo** | Mean difference | 95% CI of the difference | p-value | Mean difference | 95% CI of the difference | p-value | Mean difference | 95% CI of the difference | p-value | p-value for treatment |
| Systolic blood pressure (mmHg) | -2.5 | -14.5 to 9.5 | 0.6804 | -2.0 | -18.3 to 14.3 | 0.8037 | 10.7 | -2.2 to 23.7 | 0.1027 | 0.6084 |
| Diastolic blood pressure (mmHg) | -1.0 | -7.6 to 5.6 | 0.7727 | -2.5 | -11.5 to 6.4 | 0.5750 | 3.8 | -3.3 to 10.9 | 0.2847 | 0.9572 |
| Glycoprotein acetyls (mM) | 0.04 | -0.01 to 0.10 | 0.1002 | -0.003 | -0.07 to 0.07 | 0.9418 | -0.028 | -0.08 to 0.03 | 0.3241 | 0.1753 |
| Blood glucose (mg/dL) | 10.2 | 0.9 to 19.5 | 0.0313 | -3.7 | -16.2 to 8.7 | 0.5478 | 1.1 | -8.6 to 11.0 | 0.8278 | 0.4150 |
| Blood cholesterol (mM) | -0.07 | -0.45 to 0.30 | 0.7110 | -0.12 | -0.63 to 0.40 | 0.6506 | -0.09 | -0.50 to 0.31 | 0.6508 | 0.4622 |
| HDL-cholesterol (mM) | -0.06 | -0.15 to 0.03 | 0.2157 | 0.01 | -0.12 to 0.14 | 0.9000 | 0.02 | -0.08 to 0.12 | 0.7192 | 0.7308 |
| LDL-cholesterol (mM) | -0.02 | -0.22 to 0.17 | 0.7984 | -0.05 | -0.32 to 0.22 | 0.6962 | -0.07 | -0.29 to 0.14 | 0.4940 | 0.4472 |
| Triacylglycerides (mM) | 0.09 | -0.15 to 0.34 | 0.4388 | -0.01 | -0.34 to 0.31 | 0.9261 | -0.19 | -0.45 to 0.06 | 0.1427 | 0.6384 |
| LP-IR score | 0.37 | -2.57 to 3.3 | 0.7976 | -0.44 | -2.45 to 1.57 | 0.6455 | 0.28 | -0.95 to 1.52 | 0.6400 | 0.8888 |

## 4. Discussion

The present study evaluated the effect of the supplementation with an optimized aged black garlic (ABG) extract on the lipoprotein profile of subjects with Grade I hypertension undergoing pharmacological treatment for hypertension. Since 1993, a total of 16 meta-analyses has been conducted on the effect of garlic consumption on lipid profiles. The results have varied, with some studies demonstrating significant reductions in cholesterol levels (Ried, 2016), while others have observed no effect (Khoo & Aziz, 2009). The main limitations of these meta-analyses are the lack of information regarding the type of garlic used, as well as the standardization of its content and bioactive compounds. Most studies suggest that effectiveness may be limited to a specific population group, particularly older adults and individuals with hyperlipidemia (Varade et al., 2024). Likewise, effective doses are estimated to range between 500 and 1000 mg of garlic per day, doses that are not well tolerated by many individuals.

The importance of this study lies in the fact that it provides evidence on ABG in the lipid profile, for which there is currently limited information (only three clinical trials). Two of these trials use high doses of ABG (6 and 12 g per day) (Jung et al., 2014; Villaño et al., 2023), while one employs the same dose used in this study (Valls et al., 2022). In all three studies mentioned, no significant changes in the blood lipid profile were observed. The present study provides relevant information on an ABG extract with a low and standardized dose of S-allylcysteine (0.25 mg/day) which was highly tolerated by study participants, as well as the observation of its effects on the blood lipoprotein profile subclasses. To this regard, detailed analyses of lipoprotein subclasses showed a reduction in total lipoprotein particles, which likely reflects a lower number of atherogenic particles and, consequently, a reduced cardiovascular risk (von Eckardstein et al., 2023). Although HDL particle numbers also decreased, this was accompanied by favorable compositional changes, including higher phospholipid and lower triglyceride content in large HDL, suggesting improved HDL functionality (Chen et al., 2024). In addition, the remodeling observed in XXL-VLDL particles, characterized by higher triglyceride and lower cholesterol content, may indicate a shift toward less cholesterol-rich remnants, which are known to contribute to atherosclerosis (Ginsberg et al., 2021). The complementary changes in large HDL composition are consistent with more mature and functionally active HDL particles that may better support reverse cholesterol transport (Pownall et al., 2021). Overall, these findings suggest a beneficial remodeling of lipoproteins toward a less atherogenic profile and support the possibility that ABG supplementation can modulate lipid metabolism even at low doses.

Consistent with prior evidence, the present study shows that longitudinal changes in cholesterol levels track closely with concomitant changes in blood pressure (Huang et al., 2024; Vazquez-Agra et al., 2024). Against this background, it was observed that ABG treatment attenuated the positive coupling between temporal trajectories of cholesterol and systolic blood pressure suggests that this intervention may disrupt shared pathophysiological pathways linking dyslipidaemia to vascular dysfunction and hypertension.

Similarly to previous studies on the impact of ABG on lipid profiles, our findings indicate that the observed modifications in lipoprotein composition may depend on the individual’s baseline metabolic status. The cluster analysis performed identified that subgroups of patients with different metabolic characteristics presented varied responses to the intervention. Notably, participants classified in Cluster 3, characterized by a more adverse metabolic profile, showed a significant reduction in triglyceride levels after ABG supplementation. This finding is relevant since elevated triglyceride levels in very-low-density lipoprotein (VLDL) fractions are associated with increased cardiovascular risk (Nordestgaard, 2016). These findings highlight the importance of considering metabolic heterogeneity in the design of future investigations on the effects of black garlic on cardiovascular health.

This study has several limitations that we consider should be contemplated for further studies with the aim to validate the lipid lowering effect of ABG extracts. First, it seems that the main effects may be observed in patients with impaired lipid control and metabolism. Therefore, further studies should include as the main inclusion criteria subjects with hypercholesterolemia and more than three components of the metabolic syndrome classification according to the NCEP ATPIII. Regarding the inclusion of volunteers with lipid-lowering therapy, our data suggests that no differences in treatment were observed due to drug therapy. However, safety consideration should be taken since garlic may improve statin half-life and side-effects (Dilip Reddy et al., 2012; Reddy et al., 2010)

## 5. Conclusions

It was concluded that the supplementation with an optimized aged black garlic extract for 12 weeks in patients with Grade I hypertension modulate the number and composition of lipoprotein subclasses associated to metabolic syndorme. To this regard, cluster analysis suggests that treatment response may be influenced by participants’ baseline metabolic profile, where volunteers with impaired lipid profile may observed a reduction in TG. This suggests that ABG could be a valuable adjunct to conventional therapies for managing lipid profiles and related metabolic risks in this population. Further research is warranted to explore the underlying mechanisms and identify specific patient subgroups who may benefit most from ABG.

## Data Availability

The data that supports the findings of this study are available from the corresponding author, J CE Serrano, upon reasonable request.

## Acknowledgements

We thank the personnel of the Biobank and Pharmacology units of the IRBLleida research institute for assistance with handling human samples.

## Author Contributions

CRediT author statement: J.C.E.S.: Conceptualization, Methodology, Validation, Formal analysis, Writing, Review and Editing. E.C.-B.: Investigation. A.G.-C.: Investigation. M.I.M.-V.: Funding acquisition, Resources, Review and Editing. M.D.M.: Funding acquisition, Resources, Review and Editing. M.B.-L.: Investigation. J.M.V.: Methodology, Resources. A.E.E.: Methodology, Resources, Funding Acquisition, Review and Editing. M.P.-O.: Conceptualization, Methodology, Validation, Writing, Review and Editing. All authors have read and agreed to the published version of the manuscript.

## Declaration of generative AI and AI-assisted technologies in the manuscript preparation process

During the preparation of the manuscript COPILOT was used to correct grammar, spelling, etc. After using it, all authors reviewed and takes full responsibility for the content of the published article.

