## Supplementary material for "Aged-black garlic (Abg10+®) supplementation effect in blood lipoprotein profile in Grade I Hypertensive subjects. A randomized, triple-blind controlled trial": Suplemmentary Tables

**Figure S1.** Forest plot illustrating differences in lipoprotein particles between treatments. Point estimates the differences between Placebo and ABG group and the respective 95% interval of confidence of the difference. Robustness of significance is indicated by color: red indicates p-value <0.05; green indicates p-value <0.01; and purple indicates p-value <0.0001

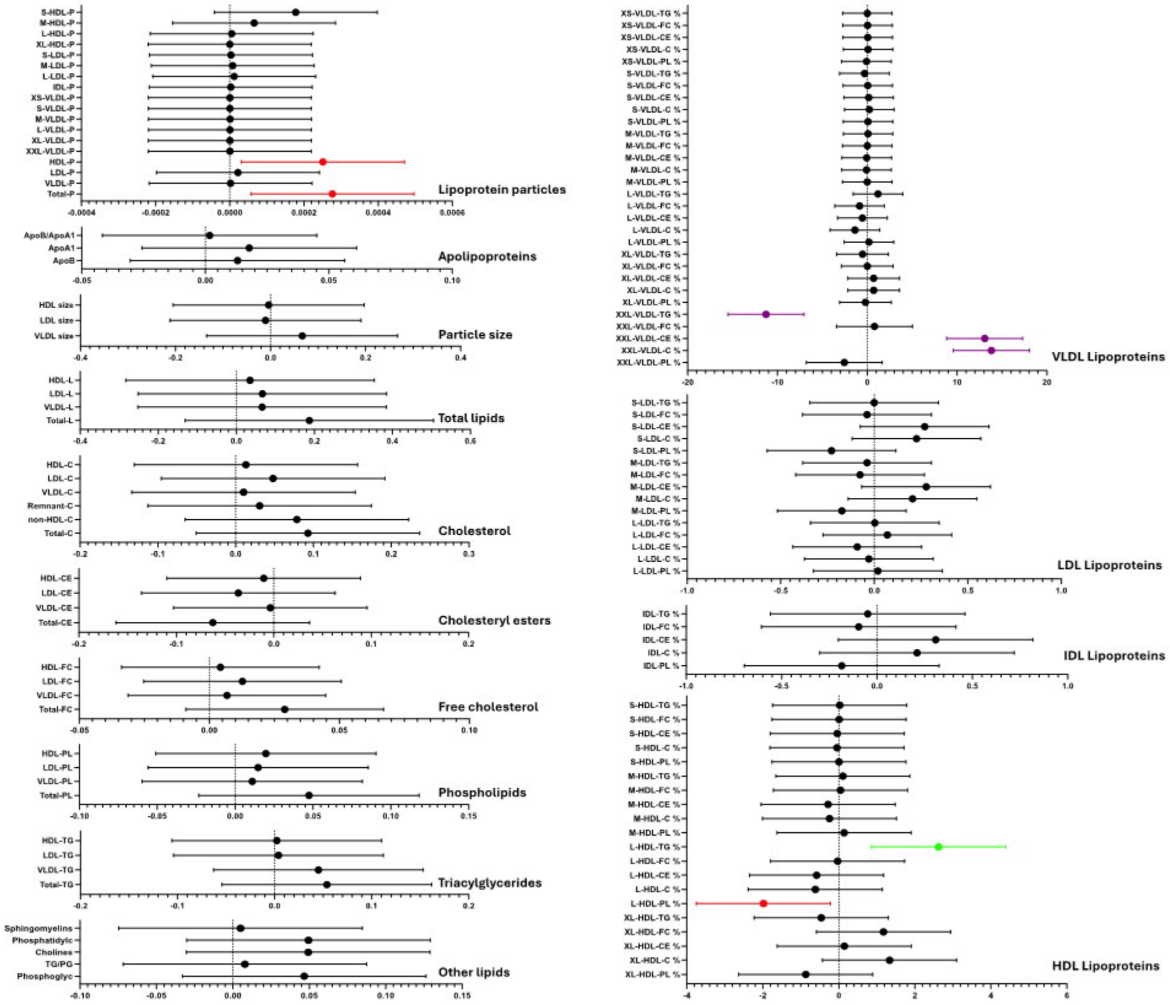

**Table S1**. Changes in lipoprotein subclasses classified by sex and treatment. Data is presented as mean ± standard deviation of the difference between 12 weeks treatment and basal conditions. Differences between groups was determined by two-way ANOVA. Bonferroni's multiple comparison test was used to determine differences by time and group treatment. P-value below 0.05 were considered as statistical differences between groups.

| Parameter | Placebo female (n=16) | Placebo male (n=20) | p-value | ABG female (n=19) | ABG male (n=20) | p-value | Placebo-ABG Female p-value | Placebo-ABG Male p-value |
|---|---|---|---|---|---|---|---|---|
| Total cholesterol mM | 0.268±0.450 | 0.033±0.404 | 0.0063 | 0.177±0.600 | -0.080±0.520 | 0.0499 | 0.9915 | 0.8273 |
| Non-HDL-cholesterol mM | 0.194±0.394 | 0.043±0.375 | 0.2858 | 0.139±0.547 | -0.070±0.444 | 0.2207 | >0.9999 | 0.8265 |
| Remnant-cholesterol mM | 0.082±0.165 | 0.022±0.162 | 0.9983 | 0.047±0.241 | -0.010±0.207 | >0.9999 | >0.9999 | >0.9999 |
| VLDL-cholesterol mM | 0.029±0.094 | $-3.0x10^{-4}$±0.096 | >0.9999 | 0.012±0.132 | -0.007±0.111 | >0.9999 | >0.9999 | >0.9999 |
| LDL-cholesterol mM | 0.112±0.241 | 0.021±0.228 | 0.9291 | 0.093±0.320 | -0.061±0.251 | 0.6993 | >0.9999 | 0.9850 |
| HDL-cholesterol mM | 0.074±0.143 | -0.010±0.107 | 0.9622 | 0.037±0.144 | -0.010±0.117 | >0.9999 | >0.9999 | >0.9999 |
| Total triacylglycerides mM | 0.066±0.244 | -0.032±0.264 | 0.8824 | -0.079±0.417 | -0.006±0.427 | 0.9993 | 0.7408 | >0.9999 |
| VLDL-triacylglycerides mM | 0.041±0.202 | -0.031±0.238 | 0.9905 | -0.074±0.380 | 0.016±0.401 | >0.9999 | 0.9419 | >0.9999 |
| LDL-triacylglycerides mM | 0.009±0.023 | 0.002±0.018 | >0.9999 | -0.001±0.016 | 0.003±0.024 | >0.9999 | >0.9999 | >0.9999 |
| HDL-triacylglycerides mM | 0.011±0.027 | -0.004±0.023 | >0.9999 | -0.004±0.023 | 0.004±0.028 | >0.9999 | >0.9999 | >0.9999 |
| Total particles mM | 0.001±0.001 | $-1.0x10^{-4}$±0.001 | >0.9999 | $4.0x10^{-4}$±0.001 | $-2.1x10^{-4}$±0.001 | >0.9999 | >0.9999 | >0.9999 |
| VLDL particles mM | $6.2x10^{-6}$±$2.1x10^{-5}$ | $-9.8x10^{-7}$±$2.0x10^{-5}$ | >0.9999 | $2.6x10^{-9}$±$2.9x10^{-5}$ | $4.8x10^{-7}$±$2.5x10^{-5}$ | >0.9999 | >0.9999 | >0.9999 |
| LDL particles mM | $5.0x10^{-5}$±$1.3x10^{-4}$ | $1.8x10^{-5}$±$1.3x10^{-4}$ | >0.9999 | $4.6x10^{-5}$±$1.8x10^{-4}$ | $-2.4x10^{-5}$±$1.4x10^{-4}$ | >0.9999 | >0.9999 | >0.9999 |
| HDL particles mM | 0.001±0.001 | $-1.3x10^{-4}$±0.001 | >0.9999 | $3.5x10^{-4}$±0.001 | $-1.9x10^{-4}$±0.001 | >0.9999 | >0.9999 | >0.9999 |

**Table S2**. Changes in lipoprotein subclasses classified by volunteers with lipid-lowering therapy and treatment. Data is presented as mean ± standard deviation of the difference between 12 weeks treatment and basal conditions. Differences between groups was determined by two-way ANOVA. Bonferroni's multiple comparison test was used to determine differences by time and group treatment. P-value below 0.05 were considered as statistical differences between groups.

| Parameter | Placebo (n=21) | Placebo Lipid-lowering (n=15) | p-value | ABG (n=23) | ABG Lipid-lowering (n=16) | p-value | Placebo-ABG p-value | Placebo-ABG Lipid-lowering p-value |
|---|---|---|---|---|---|---|---|---|
| Total cholesterol mM | 0.211±0.443 | 0.035±0.419 | 0.1971 | 0.005±0.553 | 0.101±0.603 | 0.9967 | 0.1237 | >0.9999 |
| Non-HDL-cholesterol mM | 0.185±0.402 | 0.005±0.348 | 0.1748 | -0.008±0.461 | 0.089±0.565 | 0.9963 | 0.1881 | 0.9900 |
| Remnant-cholesterol mM | 0.078±0.172 | 0.008±0.147 | 0.9978 | 0.006±0.211 | 0.034±0.246 | >0.9999 | 0.9988 | >0.9999 |
| VLDL-cholesterol mM | 0.028±0.094 | -0.009±0.096 | >0.9999 | -0.006±0.120 | 0.015±0.124 | >0.9999 | >0.9999 | >0.9999 |
| LDL-cholesterol mM | 0.108±0.245 | -0.003±0.212 | 0.8663 | -0.014±0.271 | 0.055±0.328 | >0.9999 | 0.8523 | >0.9999 |
| HDL-cholesterol mM | 0.026±0.126 | 0.029±0.138 | >0.9999 | 0.014±0.157 | 0.012±0.086 | >0.9999 | >0.9999 | >0.9999 |
| Total triacylglycerides mM | 0.056±0.282 | -0.050±0.208 | 0.8948 | -0.056±0.458 | -0.020±0.367 | >0.9999 | 0.9140 | >0.9999 |
| VLDL-triacylglycerides mM | 0.053±0.240 | -0.072±0.179 | 0.7225 | -0.061±0.422 | -0.021±0.342 | >0.9999 | 0.9060 | >0.9999 |
| LDL-triacylglycerides mM | 0.003±0.022 | 0.009±0.018 | >0.9999 | $-4.4\times10^{-6}$±0.025 | 0.003±0.011 | >0.9999 | >0.9999 | >0.9999 |
| HDL-triacylglycerides mM | -0.001±0.029 | 0.006±0.021 | >0.9999 | 0.003±0.028 | -0.005±0.021 | >0.9999 | >0.9999 | >0.9999 |
| Total particles mM | $3.8\times10^{-4}$±0.001 | $3.5\times10^{-4}$±0.001 | >0.9999 | $9.5\times10^{-5}$±0.002 | $8.0\times10^{-5}$±0.001 | >0.9999 | >0.9999 | >0.9999 |
| VLDL particles mM | $4.9\times10^{-6}\pm2.2\times10^{-5}$ | $-1.5\times10^{-6}\pm1.9\times10^{-5}$ | >0.9999 | $-1.2\times10^{-6}\pm2.8\times10^{-5}$ | $2.3\times10^{-6}\pm2.5\times10^{-5}$ | >0.9999 | >0.9999 | >0.9999 |
| LDL particles mM | $5.8\times10^{-5}\pm1.3\times10^{-4}$ | $-3.8\times10^{-6}\pm1.2\times10^{-4}$ | >0.9999 | $-1.2\times10^{-5}\pm1.4\times10^{-4}$ | $4.2\times10^{-5}\pm1.8\times10^{-4}$ | >0.9999 | >0.9999 | >0.9999 |
| HDL particles mM | $3.0\times10^{-4}$±0.001 | $3.5\times10^{-4}$±0.001 | >0.9999 | $1.0\times10^{-4}$±0.001 | $2.9\times10^{-5}$±0.001 | >0.9999 | >0.9999 | >0.9999 |
